# AI-aided dynamic prediction of bleeding and ischemic risk after coronary stenting and subsequent DAPT

**DOI:** 10.1101/2022.02.05.22270508

**Authors:** Fang Li, Laila Rasmy, Yang Xiang, Jingna Feng, Jingcheng Du, David Aguilar, Abhijeet Dhoble, Qing Wang, Shuteng Niu, Xinyue Hu, Yifang Dang, Xinyuan Zhang, Ziqian Xie, Yi Nian, JianPing He, Yujia Zhou, Ahmed Abdelhameed, Jiang Bian, Degui Zhi, Cui Tao

**Affiliations:** School of Biomedical Informatics, University of Texas Health Science Center at Houston, Houston, Texas; Peng Cheng Laboratory, Shenzhen, Guangdong, China; Department of Internal Medicine, McGovern Medical School, University of Texas Health Science Center at Houston, Houston, Texas; Division of Cardiovascular Medicine, University of Kentucky, Lexington, Kentucky; Department of Health Outcomes and Biomedical Informatics, College of Medicine, University of Florida, Gainesville, Florida

**Keywords:** artificial intelligence, dual antiplatelet therapy, bleeding risk, ischemic risk, drug-eluting stent implantation, dynamic prediction

## Abstract

**Background:** Contemporary risk scores for ischemic or bleeding event prediction after drug-eluting stent (DES) implantation are limited to the determination of a single time duration for dual antiplatelet therapy (DAPT) and lack flexibility in providing dynamic risk stratification.

**Objectives:** This study sought to develop artificial intelligence (AI) models to dynamically predict the ischemic and bleeding risks at different time intervals for patients with DES implantation for personalized decision support for antiplatelet therapy.

**Methods:** We identified 81,594 adult patients who received DES implantation in the United States from the Cerner HealthFacts^®^ dataset. The total prediction window covered 12-30 months after DES implantation. We designed eight prediction scenarios with four prediction intervals (3, 6, 12, and 18 months). Five AI models were developed for the ischemic and bleeding risk stratification. Model performance was assessed using the area under the receiver operating characteristic curve (AUROC).

**Results:** Our proposed AI model outperformed the clinical guideline-recommended tool–the DAPT score– for 12m-30m prediction (with AUROC 0.82 vs. 0.79 for ischemia, 0.77 vs 0.72 for bleeding). In the scenarios that are not covered by the DAPT score, our models demonstrated robust performance (AUROC ranges were 0.79–0.80 for ischemia and 0.75–0.76 for bleeding).

**Conclusions:** As the first effort dedicated to dynamically forecasting adverse endpoints after DES implantation given DAPT continuation or discontinuation, our AI-empowered approach demonstrates superior capabilities for risk stratification, holding value as a novel clinical tool that can refine the prognostic judgments of clinicians and achieve optimal DAPT management.

**Condensed abstract:** We proposed an innovative AI-based dynamic prediction system that forecasts the ischemic and bleeding events after coronary stenting in varying time intervals given DAPT continuation or discontinuation. Our AI model not only demonstrated superiority compared with the clinical guideline-recommended tool–the DAPT score in the 12-30 months prediction, but also achieved robust performance in other scenarios that were not covered by the DAPT score. Our AI-driven approach holds value as a novel clinical tool that can refine the prognostic judgments of clinicians, enable better informed clinical decisions, and facilitate optimal DAPT management in the context of precision cardiovascular medicine.

## Introduction

Coronary artery disease (CAD) remains a leading cause of morbidity and mortality globally (1). Percutaneous coronary intervention (PCI) is among the most frequently performed procedures in the treatment of CAD (2). As an advanced form of PCI, drug-eluting stent (DES) implantation consistently reduces the risk of restenosis, but the ischemic risk remains (3). Dual antiplatelet therapy (DAPT), which combines aspirin and an oral P2Y12 receptor inhibitor (clopidogrel, ticagrelor, and prasugrel), targets the reduction of ischemic recurrences (4). Management of DAPT, however, poses a dilemma in which prolonged duration could lead to bleeding while short duration could increase the ischemic risk (5). Therefore, assessing the balance between ischemia and bleeding liability and determining the optimal DAPT duration accordingly is of paramount importance to guide clinical decision making and improve quality of care.

The cardiovascular health community has made a substantial effort to gauge the bleeding and ischemic risk in patients treated with DAPT. Among the CAD and PCI-related scores, the CRUSADE (Can Rapid Risk Stratification Of Unstable Angina Patients Suppress Adverse Outcomes With Early Implementation of the ACC/AHA Guidelines) (6) score and the PRECISE-DAPT (Predicting Bleeding Complication in Patients Undergoing Stent Implantation and Subsequent Dual Antiplatelet Therapy) score (7) focus on bleeding risk, assessing in-hospital major bleeding and out-of-hospital bleeding during DAPT (3–6 months vs. 12–24 months), respectively. The PARIS (Patterns of Non-Adherence to Antiplatelet Regimen in Stented Patients) risk score (8) and the DAPT score (9) predict both bleeding and ischemic endpoints with prolonged DAPT, with a forecasting window of 0–24 months and 12–30 months, respectively. These scores leverage a handful of features to inform clinical decisions regarding risk avoidance strategies. Nevertheless, the have certain limitations: (1) The prediction windows are unitary and static, such as 12 months after discharge (the PRECISE-DAPT score for bleeding risk estimation) or 18 months after completion of first-year DAPT without any major events (the DAPT score for ischemic and bleeding risk assessment). The static functionality is insufficient to support flexible and granular risk estimation. (2) The existing parametric regression models applied in these scores did not fully exploit the full potential of the high-dimensional variables collected in the real-world data (10). For example, the emerging predictors for bleeding might be missing during the development of the PRECISE-DAPT score. To enhance the discriminative capability of the model, the authors have clarified that they would consider clinical, laboratory, or genetic factors in future studies (7). (3) The rigid inclusion criteria in clinical trials may induce estimation bias and the resulting estimated efficacy may be different from the effectiveness of the general patient population. For instance, the PARIS risk score provides an analysis of the patients who were exclusively treated with clopidogrel, which constrains its generalizability to more potent P2Y12 inhibitors (8). Therefore, advanced decision support tools that can overcome these shortcomings and provide dynamic and personalized risk prediction are urgently needed.

Artificial intelligence (AI), coupled with promising machine learning (ML) techniques well known in computer science, is having a broad effect on various fields including science and technology, and industry, as well as in our day-to-day life (11). AI refers to the simulation of human intelligence by a system or a machine. The goal of AI researchers is to develop a machine that can think like humans and mimic human behaviors, including perceiving, reasoning, learning, planning, predicting, and so on (12). Over the past decade, AI techniques, especially ML and deep learning (DL), have been successfully applied in cardiovascular (CV) medicine–bringing tremendous precision to diagnosis and prognostication–and may play a critical role in facilitating evolution of CV medicine to precision CV medicine (12, 13). AI algorithms have been increasingly employed in the CAD-related prediction tasks, yielding prime performance compared with the classic statistical models (14–17). Contemporary studies, however, can provide only a fixed window prediction with static input data. None has developed the risk scores based on variables that become available as therapy progresses. In addition, the majority of the studies adopted a time-consuming and labor-intensive process in which researchers need to carefully handcraft features from the raw data (18). Targeting the limitations of the current static scores or traditional feature engineering-based ML models, we propose a novel non-feature selection-based approach that leverages cutting-edge AI models to dynamically predict the bleeding and ischemic risks after DES implantation, given DAPT continuation or discontinuation, by incorporating updates of clinical data.

**Central illustration.**
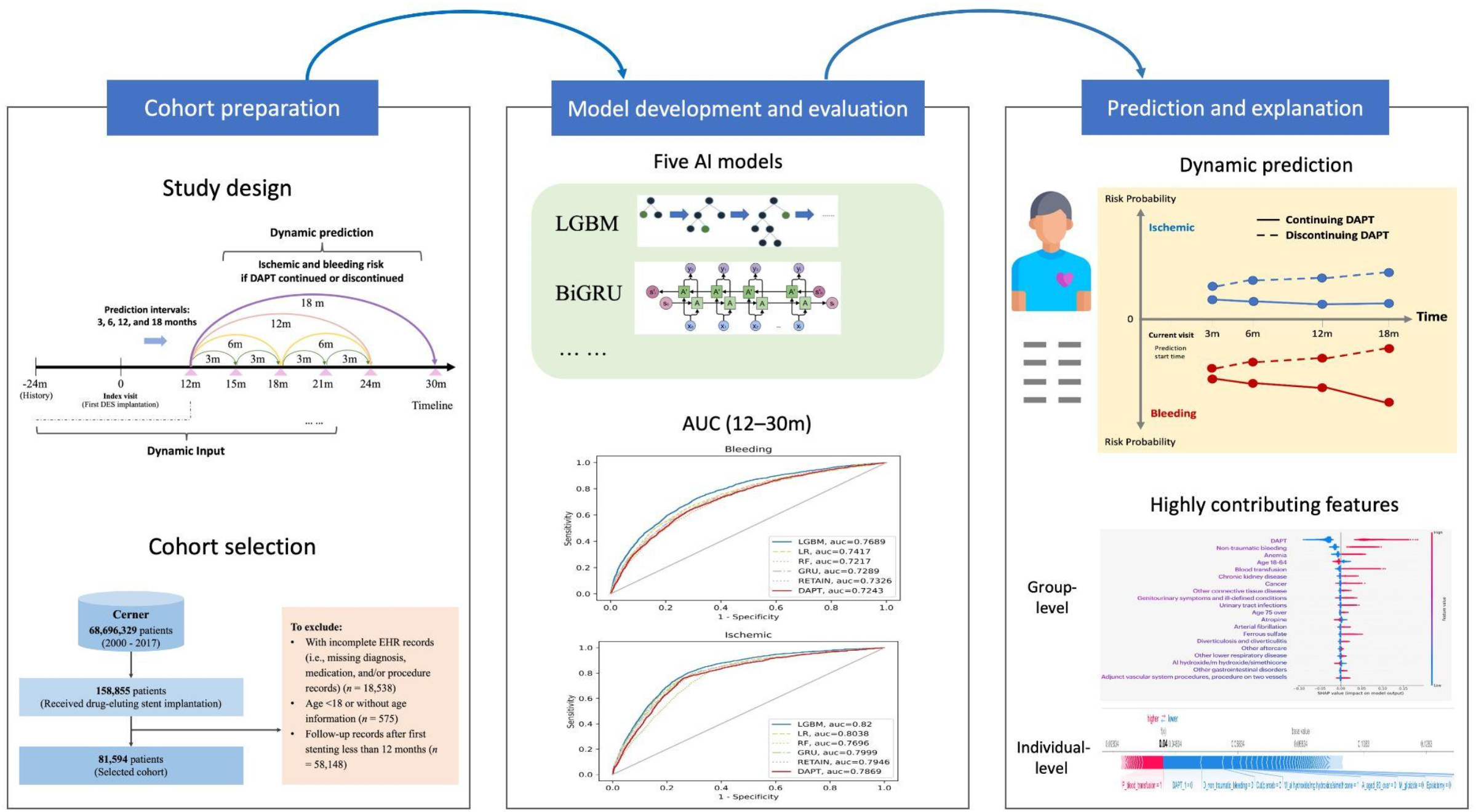
Central Illustration shows the AI-driven dynamic prediction framework, which comprises cohort preparation, model development and evaluations, and prediction and explanation.

## Methods

### Data Source and Study population

The cohort was extracted from the Cerner Health-Facts^®^ (referred to as Cerner in this study), a large real-world de-identified dataset. Cerner (2017 version) incorporates electronic health records (EHRs) from over 600 participating client hospitals and clinics in the United States, representing over 68 million unique patients from 2000 to 2017 (19). UTHealth has agreements with Cerner to use data for research purposes, and the Institutional Review Board (IRB) at UTHealth has approved the study protocol. The data comprise demographics, encounters, diagnoses, procedures, lab results, medications, and other clinical observations.

Consecutive patients, who received DES implantation between January 1, 2000 and December 31, 2017, were identified for potential inclusion in this study (*N* = 158,855) (see Supplementary Table 1 for the DES implantation codes). The index visit was defined as the visit when a patient underwent the first DES implantation. Patients were excluded if they met any of the following criteria: (1) missing critical information, including without diagnosis, medication, or procedure records (*n* = 18,538); (2) age < 18 years old or without age information (n = 575); or (3) follow-up after index visit shorter than 12 months (*n* = 58,148).

### Outcomes and Characteristics

The positive effects of DAPT are assumed to reduce both cardiac and/or cerebral ischemia and revascularization procedures. Thus, the primary ischemic endpoint was defined as a composite of acute ischemic heart disease, ischemic stroke, multiple DES implantation, and coronary artery bypass graft (CABG). The primary bleeding endpoint was a composite of minor to severe spontaneous bleeding and blood transfusion procedure, as defined by the GUSTO (the Global Use of Strategies to Open Occluded Arteries) criteria (20) (see Supplementary Tables 2 and 3 for the detailed diagnosis and procedure codes of the ischemic and bleeding endpoints). The standard terminologies referenced comprised the International Classification of Diseases (ICD) (including ICD-9-CM and ICD-10-CM with both diagnoses and procedures codes), Current Procedural Terminology, 4th Edition (CPT-4), and the Healthcare Common Procedure Coding System (HCPCS).

Patient characteristics that were potentially associated with the development of the endpoints (9, 21) were extracted and tagged from Cerner, comprising: (1) basic demographics, including the age at the index visit, sex, and race/ethnicity; (2) medical history and comorbidities, including prior non-traumatic bleeding, prior myocardial infarction (MI), stroke, alcohol abuse, smoking-related disease, anemia, atrial fibrillation, cancer, chronic kidney disease (CKD), diabetes mellitus, dyslipidemia, congestive heart failure, hypertension, liver disease, peripheral vascular disease (PVD), and prior CABG; (3) concomitant medications, including angiotensin-converting enzyme inhibitors (ACEIs), angiotensin receptor blockers (ARBs), beta-blockers, calcium antagonists, non-steroidal anti-inflammatory drugs (NSAIDs), and statins. We referenced the Elixhauser Comorbidity Index (22) for comorbidity definitions and Epocrates Drugs (23) for medication definitions (see Supplementary Tables 4 and 5 for the specific ICD codes of comorbidities and generic name sets of the comedications).

### Prediction Tasks

We hypothesized that AI models can provide optimal and personalized DAPT duration recommendations in consideration of patients’ unique characteristics, thus minimizing the risk of severe adverse events (ischemic diseases or life-threatening bleeding) after receiving DES implantation. Based on this hypothesis, our study attempted to dynamically predict patients’ ischemic and bleeding risks covering 12–30 months after first stenting with varying lengths of forecasting windows.

DAPT exposure was defined as the combinational antiplatelet therapy with aspirin and a P2Y12 receptor inhibitor (clopidogrel, prasugrel, or ticagrelor). The recommended minimum duration in patients treated with DES is 12 months, based on the 2016 ACC/AHA Guideline (24). Hence, we assumed that DAPT for the first 12 months was a default treatment. To assist decision-making for the optimum continuation, we defined 12–30 months after the first DES implantation as the total prediction range, identical to that of the DAPT score. In contrast to the static function of the DAPT score (i.e., one prediction window with a unique length of 18 months), our program can predict risk dynamically. As a demonstration, we designated four prediction intervals (i.e., 3, 6, 12, and 18 months) to conduct dynamic and variable-length forecasting. The prediction start point could be flexible, beginning as early as 12 months after the first stenting. To demonstrate typical and clinical meaningful predictions, we designed eight scenarios in total. Taking the prediction start time at 12 months after discharge as an example, the prediction windows could be 12–15m (with an interval of 3 months), 12–18m (6 months), 12–24m (12 months), and 12–30m (18 months), while the input data were the records before prediction start time, including the first 12 months and the history (i.e., within 24 months prior to the index stenting) (Figure 1). Table 1 presents all scenarios with prediction intervals, input data, and prediction start and end times. Because dynamic prediction scenarios had a variety of prediction start time points, the final cohort for each prediction scenario was further selected as those whose last records were later than the prediction start time.

**Table 1.** Specification of Exemplar Prediction Scenarios

| Prediction intervals | Input Data | Prediction windows with start and end times<sup>a</sup> |
| --- | --- | --- |
| 3 months | History<sup>b</sup> + index visit<sup>c</sup> + 12 m records | 12m - 15m |
|  | History + index visit + 15 m records | 15m - 18m |
|  | History + index visit + 18 m records | 18m - 21m |
|  | History + index visit + 21 m records | 21m - 24m |
| 6 months | History + index visit + 12 m records | 12m - 18m |
|  | History + index visit + 18 m records | 18m - 24m |
| 12 months | History + index visit + 12 m records | 12m - 24m |
| 18 months | History + index visit + 12 m records | 12m - 30m |

<sup>a</sup>The start and end time refers to the time after the index visit
<sup>b</sup>History refers to patients' records within 24 months prior to the index visit
<sup>c</sup>Index visit refers to the visit of receiving the first drug-eluting stent implantation

**Figure 1.**
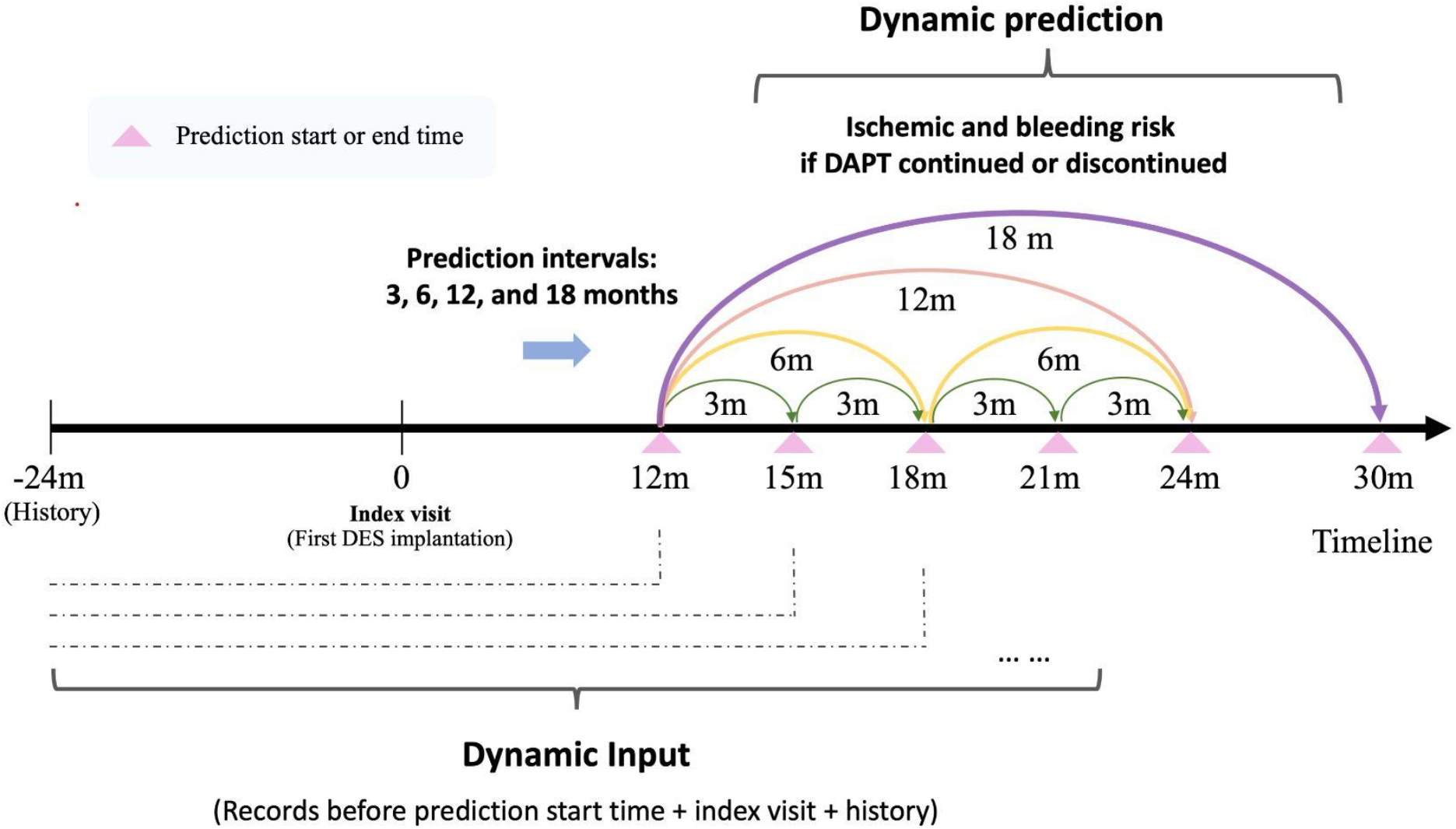
Illustration of dynamic prediction scenarios

### AI Modeling

We developed and compared five AI models for risk stratification. Three are classic ML models, including light gradient boosting machine (LGBM) (25), logistic regression (LR) (26), and random forest (RF) (27); and two are recurrent neural networks (RNN)-based DL models, including gated recurrent unit (GRU) (28) and REverse Time AttentIoN (RETAIN) (29). Specifically, LGBM is a highly efficient gradient boosting decision tree with the strengths of fast training speed and good accuracy (30). RNN is suited for predicting disease onset based on longitudinal observations of clinical events, as it takes into account the temporality in a sequence of events (31). As for the two RNN-based architectures, GRU was created to catch both short-term and long-term memory using gate mechanisms, while RETAIN aimed to improve clinical prediction modeling interpretability. RETAIN is based on a two-level sequential neural attention model that detects influential past visits and significant clinical variables within those visits. It mimics physicians’ practice by attending the EHR in reverse time order so that recent clinical visits are likely to receive higher attention (29). With respect to the implementation codes, we conducted data pre-processing on a Python-based pipeline, which resulted in patients’ comprehensive clinical profiling in a unified time sequence. We utilized the packages in scikit-learn 1.0.1 for RF and LR, an anaconda-supported package for LGBM, and an EHR-tailored predictive pipeline pytorch_ehr (32, 33) for GRU and RETAIN.

The input data comprised patients’ age, diagnoses, medications, and procedures (from two years prior to the index visit until the prediction start time) as well as DAPT continuation state in the prediction window. Based on the distribution characteristics of DATP records in Cerner, we set the DAPT continuation criterion as follows: If the patient has more than one DAPT record within 12m–30m after index stenting, he or she would be regarded as having continued DAPT; if not, then, regarded as discontinued. Using this criterion, the ratio of DAPT continuation: discontinuation within 12m–30m was around 1:2 among the cohorts (see Supplementary Table 6). The input data fed into AI algorithms were then compiled in the structure of a list, consisting of (1) unique patient ID; (2) event label, i.e., 0 for control, 1 for case; (3) information of each visit, including time gap (in days) relative to the previous visit and the medical codes (including diagnoses, medications, and procures); and (4) state label of DAPT continuation in the prediction window. To represent discrete medical codes as the computable matrix, we adopted multi-hot encodings for the classic ML models (LGBM, LR, and RF) and randomly initialized embeddings for the DL models (BiGRU and RETAIN).

We defined our prediction as single-task learning. In other words, for ischemic and bleeding events, we trained AI models for their risk stratification separately. We labeled the patients in the selected cohort as case or control for each event. Specifically, a patient was labeled as a case if he or she developed the event within the prediction window. Otherwise, the patient was labeled as control. For model development, we randomly split the cohort into training, validation, and test sets with a ratio of 7:1:2. To assist hyperparameter tuning, we utilized an open-source hyperparameter optimization framework–Optuna (34). The current state-of-the-art version (2.10.0) of Optuna was installed from Anaconda. In addition, an early stop strategy was applied for DL algorithms to speed up the training process and prevent overtraining. For discrimination evaluation, the area under the receiver operating characteristic curve (AUROC or AUC for short) analysis was used to assess the predictive capacity of AI models.

For the benchmark, two validated scores (i.e., the DAPT score and the PRECISE-DAPT score) that are extensively applied in clinical studies (35, 36), served as the potential candidates. The PRECISE-DAPT focuses mainly on forecasting the out-of-hospital bleeding risk, while the DAPT score was dedicated to estimating the incidence probabilities of both events, which matches our prediction task. Hence, we selected the DAPT score as our benchmark. In regard to methodology, the DAPT score applied multivariable Cox regression with 11 significant predictors for predictive modeling and the model discrimination was assessed using the C-statistic (8). To calculate the DAPT score, we fed the Cox model with the values of these predictors derived from the Cerner cohort. Further, we converted C-statistic to AUC to achieve a fair comparison based on the same evaluation metric (see Supplementary Appendix Heading 6–Conversion of Cox C-statistic to AUC for the details).

### Statistical Analysis and Model Interpretability

For descriptive analysis of demographic and clinical characteristics, categorical variables are reported as count (%) and continuous variables as mean (standard deviation, SD). We used the chi-square test for categorical variables and the Kruskal-Wallis test for continuous variables to assess the differences between patients who experienced events and those who did not in 12m–30m. A two-sided p < 0.05 was considered statistically significant.

Interpretability is one of the most dominant features of predictive models. We leveraged SHapley Additive exPlanations (SHAP) (37) for visualization of the vital clinical features. SHAP is a unified framework that uses the classic Shapley values to explain the output of a given model and interpret predictions (38). Specifically, Shapley values provide a calculation of the importance of a feature by comparing what a model predicts with or without the feature (39). To increase concept granularity of input data and enhance the visualization effect, we adopted a hybrid strategy for medical code conversion: (1) For age, the continuous values were converted to the categorical value, including ages 18–64, 65–74, and 75 and over 75, following the age stratification criteria of the DAPT score documented in the 2016 ACC/AHA Guideline (24); (2) For diagnoses, if the ICD codes belonged to the predefined characteristics (e.g., ischemic heart disease, congestive heart failure), we used characteristic name instead of the original code as input; if not, we converted the original ICD codes (both ICD-9-CM and ICD-10-CM) to the Clinical Classifications Software (CCS) (40). The CCS is an ICD-9-CM-based diagnosis and procedure categorization scheme. It condenses ICD-9-CM’s multitude of codes–over 14,000 diagnosis codes and 3,900 procedure codes–into a smaller number of clinically meaningful categories, which are sometimes more useful for presenting descriptive statistics than are individual ICD-9-CM codes (41); (3) For medications, if the generic name belonged to the predefined comedication feature (e.g., statins), we used predefined comedication feature name; if not, we used the original generic name.

## Results

### Baseline Characteristics

A total of 81,594 adult patients who underwent DES implantation with longer than one-year record after first stenting were included for AI model development (Figure 2). From 12 through 30 months after their index DES procedure, 11,039 (13.5%) patients developed ischemic events and 7,402 (9.1%) developed bleeding events, while 2,158 (2.6%) had both ischemic and bleeding events. For the ischemic events, with respect to demographics, patients who developed events in follow-up had a higher rate of females (36.9% vs. 35.3%, *p* = 0.002), a higher percentage of African Americans (12.7% vs. 9.2%, *p* < 0.001), and a lower percentage of Caucasians (80.3% vs. 84.1%, *p* < 0.001), compared with those without events. In terms of medical history and comorbidities, ischemic individuals had (1) a higher prevalence of cardiovascular and hematologic diseases, including prior acute myocardial infarction, prior spontaneous bleeding, hypertension, congestive heart failure, atrial fibrillation, stroke, and anemia (all *p* values < 0.001); (2) higher rates of possible risk factors and comorbidities, including diabetes, dyslipidemia, chronic kidney disease, prior smoking, alcohol abuse, liver disease, and peripheral vascular disease (all *p* values < 0.001); and higher rates of comedications, including the use of ACEIs, ARBs, beta-blockers, calcium antagonists, NSAIDs, and statins (all *p* values < 0.001). Regarding age, CABG, and cancer, no significant difference is shown between the event and the non-event group (Table 2).

**Table 2.** Baseline Characteristics of Patients With vs Without Ischemic or Bleeding Events from 12 to 30 months in the Selected Cohort (*N* = 81,594)

|  |  | Ischemic Events, No. (%) |  |  | Bleeding Events, No. (%) |  |  |
| --- | --- | --- | --- | --- | --- | --- | --- |
|  |  | Event,<br>11,039 (13.5%) | No Event,<br>70,555<br>(86.47%) | p Value | Event,<br>7,402 (9.1%) | No Event,<br>74,192 (90.9%) | p Value |
| **Demographics** |  |  |  |  |  |  |  |
| Age, Mean (SD), yrs |  | 64.3 (12.1) | 64.2 (12.0) | 0.891 | 67.1 (12.0) | 64.0 (12.0) | <0.001* |
| Female |  | 4,070 (36.9%) | 24,926<br>(35.3%) | 0.002* | 2,945 (39.8%) | 26,051 (35.1%) | <0.001* |
| Race | Caucasian | 8,866 (80.3%) | 59,326<br>(84.1%) | <0.001* | 5,991 (80.9%) | 62,201 (83.8%) | <0.001* |
|  | African American | 1,400 (12.7%) | 6,503 (9.2%) | <0.001* | 916 (12.4%) | 6,987 (9.4%) | <0.001* |
|  | Asian | 139 (1.3%) | 1,038 (1.5%) | 0.090 | 112 (1.5%) | 1,065 (1.4%) | 0.629 |
|  | Hispanic | 76 (0.7%) | 413 (0.6%) | 0.215 | 43 (0.6%) | 446 (0.6%) | 0.892 |
|  | Other<sup>a</sup> | 558 (5.1%) | 3,275 (4.6%) | 0.060 | 340 (4.6%) | 3,493 (4.7%) | 0.677 |

| Medical History and Comorbidities |  |  |  |  |  |  |
| --- | --- | --- | --- | --- | --- | --- |
| Prior bleeding | 1,872 (17.0%) | 8,875 (12.6%) | <0.001* | 2,264 (30.6%) | 8,483 (11.4%) | <0.001* |
| Prior MI | 4,729 (42.8%) | 17,878 (25.3%) | <0.001* | 2,529 (34.2%) | 20,078 (27.1%) | <0.001* |
| Stroke | 1,530 (13.9%) | 4,409 (6.2%) | <0.001* | 909 (12.3%) | 5,030 (6.8%) | <0.001* |
| CABG | 212 (1.9%) | 1,134 (1.6%) | 0.018 | 140 (1.9%) | 1,206 (1.6%) | 0.096 |
| Alcohol abuse | 404 (3.7%) | 1,803 (2.6%) | <0.001* | 327 (4.4%) | 1,880 (2.5%) | <0.001* |
| Anemia | 2,984 (27.0%) | 12,967 (18.4%) | <0.001* | 2,878 (38.9%) | 13,073 (17.6%) | <0.001* |
| Atrial fibrillation | 1,543 (14.0%) | 7,842 (11.1%) | <0.001* | 1,460 (19.7%) | 7,925 (10.7%) | <0.001* |
| Cancer | 1,319 (11.9%) | 8,241 (11.7%) | 0.424 | 1,442 (19.5%) | 8,118 (10.9%) | <0.001* |
| CKD | 2,295 (20.8%) | 8,617 (12.2%) | <0.001* | 1,979 (26.7%) | 8,933 (12.0%) | <0.001* |
| Diabetes | 4,891 (44.3%) | 22,999 (32.6%) | <0.001* | 3,506 (47.4%) | 24,384 (32.9%) | <0.001* |
| Dyslipidemia | 7,953 (72.0%) | 43,876 (62.2%) | <0.001* | 5,472 (73.9%) | 46,357 (62.5%) | <0.001* |
| Congestive heart failure | 3,526 (31.9%) | 14,346 (20.3%) | <0.001* | 2,743 (37.1%) | 15,129 (20.4%) | <0.001* |
| Hypertension | 8,283 (75.0%) | 44,580 (63.2%) | <0.001* | 5,849 (79.0%) | 47,014 (63.4%) | <0.001* |
| Liver disease | 702 (6.4%) | 3,467 (4.9%) | <0.001* | 686 (9.3%) | 3,483 (4.7%) | <0.001* |
| PAD | 3,344 (30.3%) | 14,684 (20.8%) | <0.001* | 2,523 (34.1%) | 15,505 (20.9%) | <0.001* |
| Prior Smoking | 2,374 (21.5%) | 10,591 (15.0%) | <0.001* | 1,687 (22.8%) | 11,278 (15.2%) | <0.001* |

| Comedications |  |  |  |  |  |  |
| --- | --- | --- | --- | --- | --- | --- |
| ACE Inhibitors use | 3,959 (35.9%) | 17,664 (25.0%) | <0.001* | 2,733 (36.9%) | 18,890 (25.5%) | <0.001* |
| ARBs use | 1,429 (12.9%) | 6,606 (9.4%) | <0.001* | 1,131 (15.3%) | 6,904 (9.3%) | <0.001* |
| Beta-blockers use | 5,964 (54.0%) | 28,718 (40.7%) | <0.001* | 4,278 (57.8%) | 30,404 (41.0%) | <0.001* |
| Calcium antagonists use | 3,245 (29.4%) | 14,068 (19.9%) | <0.001* | 2,386 (32.2%) | 14,927 (20.1%) | <0.001* |
| NSAIDs use | 2,234 (20.2%) | 11,482 (16.3%) | <0.001* | 1,734 (23.4%) | 11,982 (16.1%) | <0.001* |
| Statins use | 5,600 (50.7%) | 26,734 (37.9%) | <0.001* | 3,967 (53.6%) | 28,367 (38.2%) | <0.001* |

Abbreviations: ACEIs, angiotensin-converting enzyme inhibitors; ARBs, angiotensin receptor blockers; CABG, coronary bypass artery graft; CKD, chronic kidney disease; MI, myocardial infarction; NSAIDs, non-steroidal anti-inflammatory drugs; PVD, peripheral vascular disease
\* $p < 0.05$ regarded as significance. $p$ -values were estimated by the chi-square test for categorical variables and the Kruskal-Wallis test for continuous variables.
<sup>a</sup>Other group in Race includes Native American, Pacific Islander, Asian/Pacific Islander, Biracial, Mid-Eastern Indian, Other, and Unknown.

**Figure 2.**
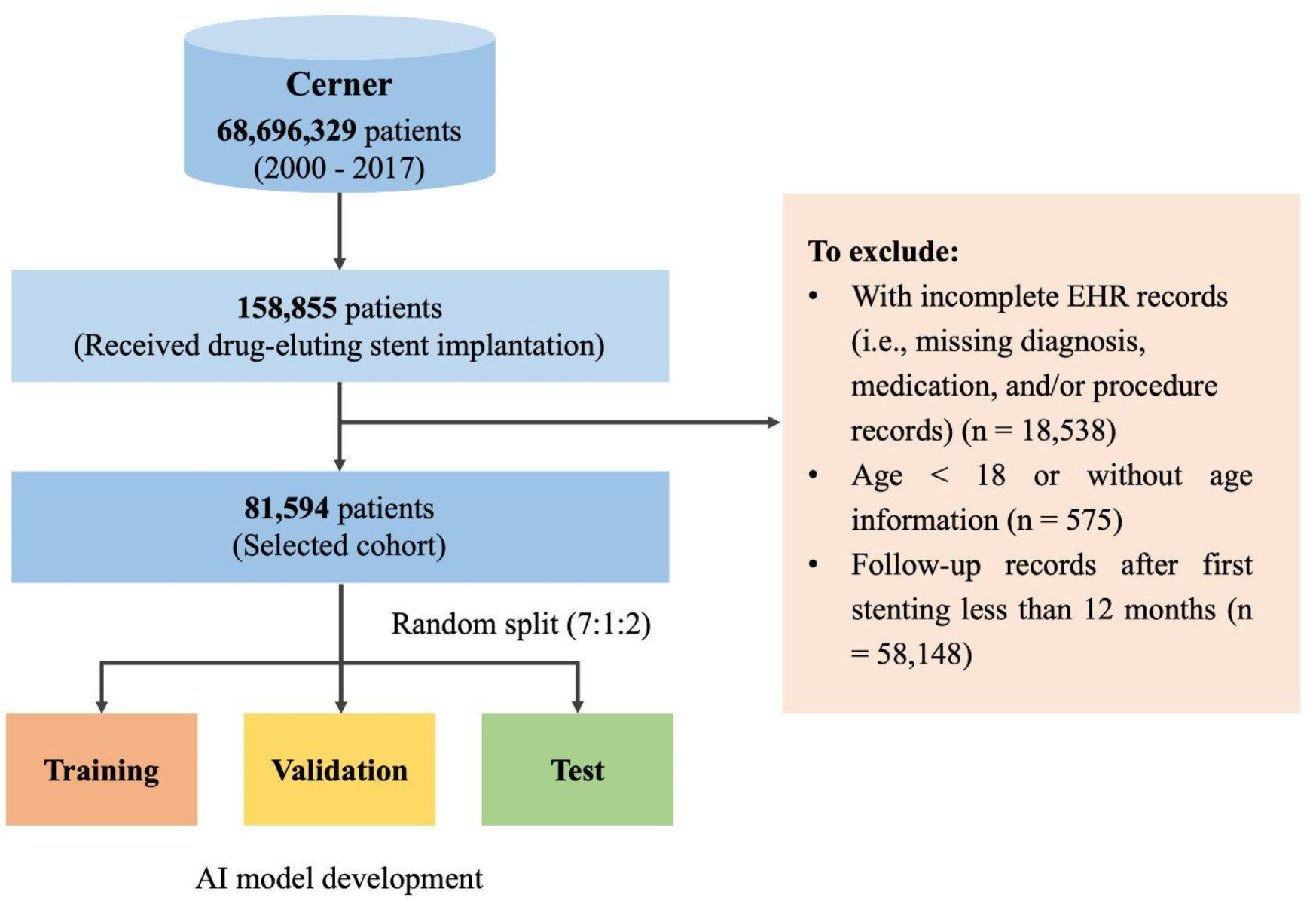
Cohort selection A total of 81,594 patients were included in the final analysis and randomly split into training, validation, and test set with a ratio of 7:1:2 for AI model development.

For the bleeding event, interestingly, patients with events shared the most characteristics with the patients who developed ischemic events. Differences were found, however, in terms of their being older (67.1 ± 12.0 vs. 64.0 ± 12.0, *p* < 0.001) and having a higher prevalence of cancer (19.5% vs. 10.9%, *p* < 0.001) as compared with the non-bleeding event group (Table 2).

### Model Performance

For ischemia forecasting, the best performance model was from LGBM, with an AUC of 0.820 in the prediction window of 12m–30m. By contrast, the DAPT score yielded a C-statistic of 0.7694, which corresponded to an AUC of 0.7869 after conversion. LGBM showed 3.3% superiority over the DAPT score. The other three AI models (BiGRU, RETAIN, and LR) also outperformed the DAPT score in this prediction task (see Figure 3a). In the other seven scenarios that were not covered by the DAPT score, LGBM also showed the best performance among the five AI models, with stable AUC scores that ranged from 0.7849 to 0.8033 (Table 3).

**Table 3.**
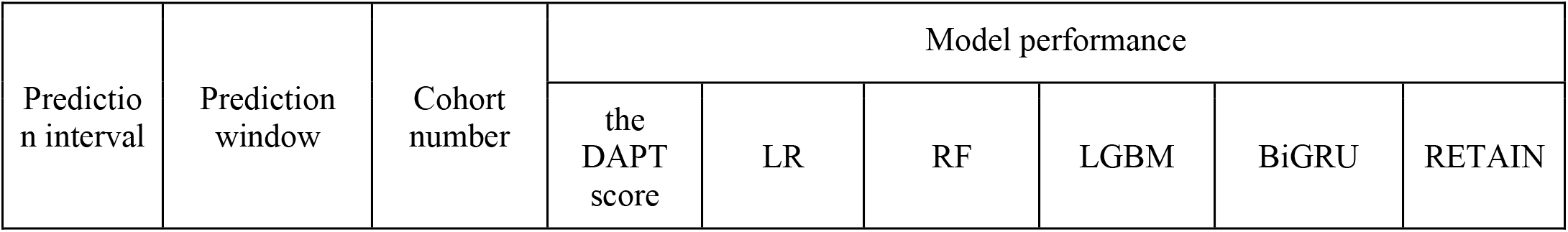
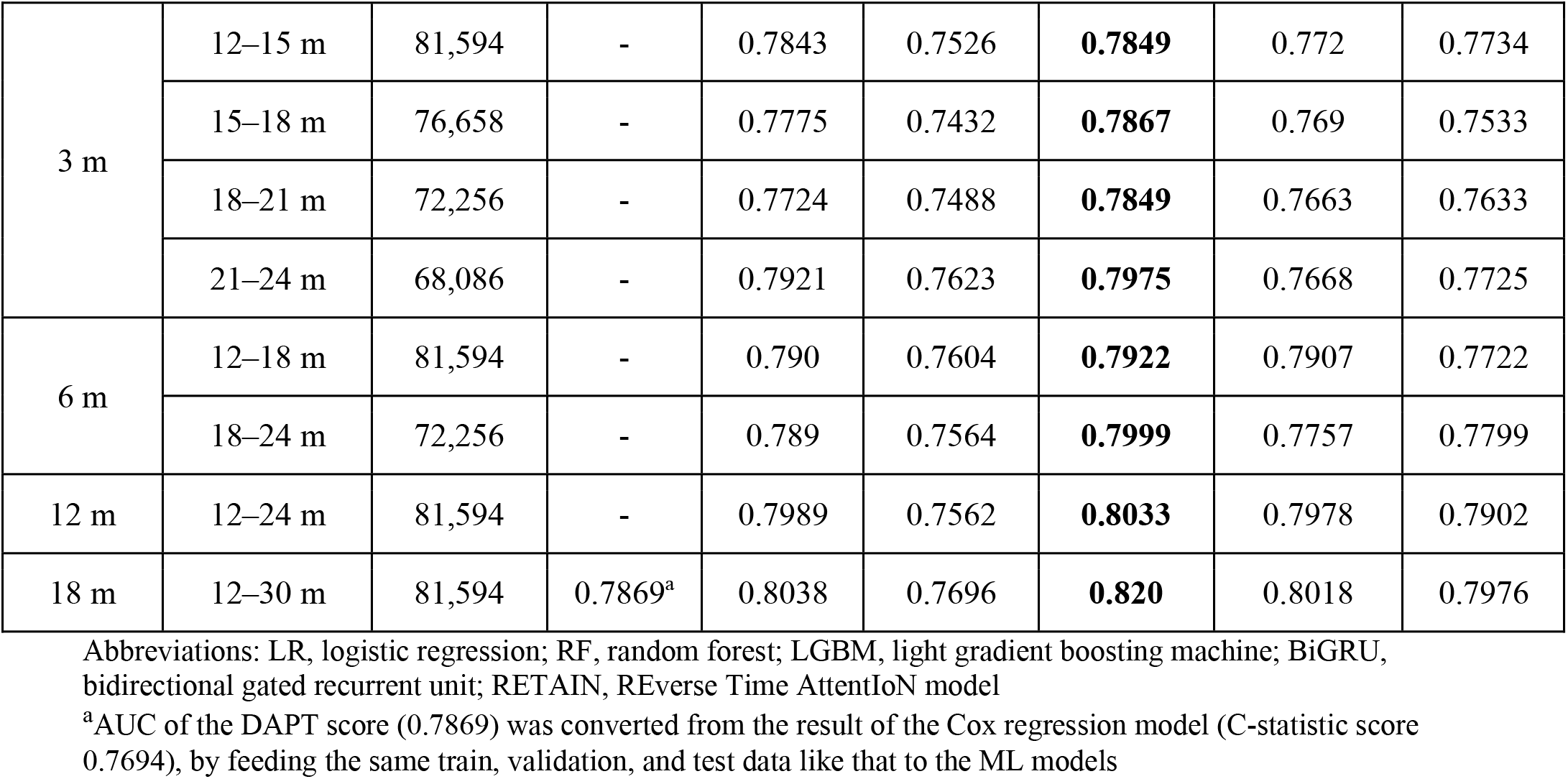
AUC for the ischemic event

| Prediction interval | Prediction window | Cohort number | Model performance |  |  |  |  |  |
| --- | --- | --- | --- | --- | --- | --- | --- | --- |
|  |  |  | the DAPT score | LR | RF | LGBM | BiGRU | RETAIN |
| 3 m | 12–15 m | 81,594 | - | 0.7843 | 0.7526 | **0.7849** | 0.772 | 0.7734 |
|  | 15–18 m | 76,658 | - | 0.7775 | 0.7432 | **0.7867** | 0.769 | 0.7533 |
|  | 18–21 m | 72,256 | - | 0.7724 | 0.7488 | **0.7849** | 0.7663 | 0.7633 |
|  | 21–24 m | 68,086 | - | 0.7921 | 0.7623 | **0.7975** | 0.7668 | 0.7725 |
| 6 m | 12–18 m | 81,594 | - | 0.790 | 0.7604 | **0.7922** | 0.7907 | 0.7722 |
|  | 18–24 m | 72,256 | - | 0.789 | 0.7564 | **0.7999** | 0.7757 | 0.7799 |
| 12 m | 12–24 m | 81,594 | - | 0.7989 | 0.7562 | **0.8033** | 0.7978 | 0.7902 |
| 18 m | 12–30 m | 81,594 | 0.7869<sup>a</sup> | 0.8038 | 0.7696 | **0.820** | 0.8018 | 0.7976 |

Abbreviations: LR, logistic regression; RF, random forest; LGBM, light gradient boosting machine; BiGRU, bidirectional gated recurrent unit; RETAIN, REverse Time AttentIoN model
<sup>a</sup>AUC of the DAPT score (0.7869) was converted from the result of the Cox regression model (C-statistic score 0.7694), by feeding the same train, validation, and test data like that to the ML models

**Figure 3.**
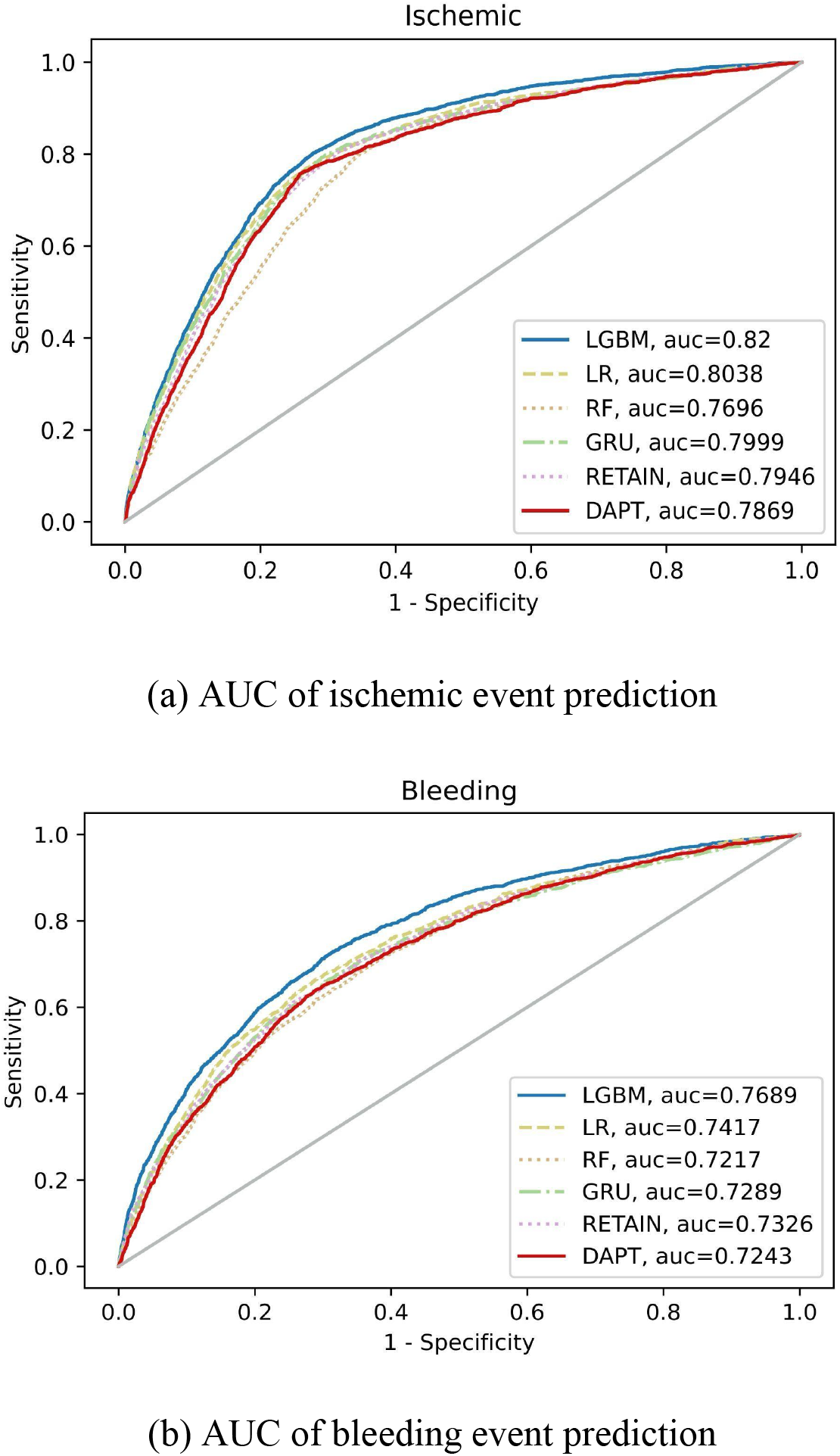
AUC of the AI models vs. the DAPT score in the prediction of ischemic and bleeding risks in the 12m–30m window

For bleeding risk assessment, LGBM demonstrated the best prediction capability as well. In the window of 12m–30m, LGBM yielded an AUC of 0.7689, outperforming the DAPT score with an AUC of 0.7243 (converted from C-statistic 0.7198). In addition, the other three AI models (BiGRU, RETAIN, and LR) also were superior to the DAPT score (See Figure 3b). In the other seven scenarios, LGBM achieved stable performance with AUC that ranged from 0.746 to 0.763 (Table 4).

**Table 4.** AUC for the bleeding event

| Prediction interval | Prediction window | Cohort number | Model performance |  |  |  |  |  |
| --- | --- | --- | --- | --- | --- | --- | --- | --- |
|  |  |  | DAPT score | LR | RF | LGBM | BiGRU | RETAIN |
| 3 m | 12–15 m | 81,594 | - | 0.750 | 0.7394 | **0.7567** | 0.7315 | 0.7121 |
|  | 15–18 m | 76,658 | - | 0.754 | 0.7402 | **0.7604** | 0.7476 | 0.7168 |
|  | 18–21 m | 72,256 | - | 0.7639 | 0.749 | **0.760** | 0.7636 | 0.7061 |
|  | 21–24 m | 68,086 | - | 0.7638 | 0.7459 | **0.763** | 0.7586 | 0.7288 |
| 6 m | 12–18 m | 81,594 | - | 0.7397 | 0.7262 | **0.746** | 0.7332 | 0.7157 |
|  | 18–24 m | 72,256 | - | 0.7609 | 0.7303 | **0.7572** | 0.7488 | 0.7485 |
| 12 m | 12–24 m | 81,594 | - | 0.7462 | 0.7227 | **0.752** | 0.7354 | 0.7242 |
| 18 m | 12–30 m | 81,594 | 0.7243<sup>a</sup> | 0.7417 | 0.7217 | **0.7689** | 0.7325 | 0.730 |

Abbreviations: LR, logistic regression; RF, random forest; LGBM, light gradient boosting machine; BiGRU, Bidirectional gated recurrent unit; RETAIN, REverse Time Attention model
<sup>a</sup>AUC of the DAPT score (0.7243) was converted from the result of the Cox regression model (C-statistic score 0.7198), by feeding the same train, validation, and test data like that to the ML models

Overall, our best AI model outperformed the DAPT score in ischemic and bleeding event prediction, while manifesting robust performance in other scenarios that were not covered by the DAPT score, regardless of the prediction intervals or windows.

### Model Explanation

To pinpoint the important features that had an impact on composite bleeding and ischemic risk prediction, we calculated the SHAP feature importance values from the input data (including both original medical codes and feature labels). We chose the 12m–30m window as the example scenario and employed LGBM, which showed the best performance among five AI models, to generate SHAP beeswarm diagrams on the population level (Figure 4). The horizontal axis represents the SHAP value, i.e., the impact on the model output of the feature, and the vertical axis (feature value) indicates the name of the features in the order of importance from top to bottom.

**Figure 4.**
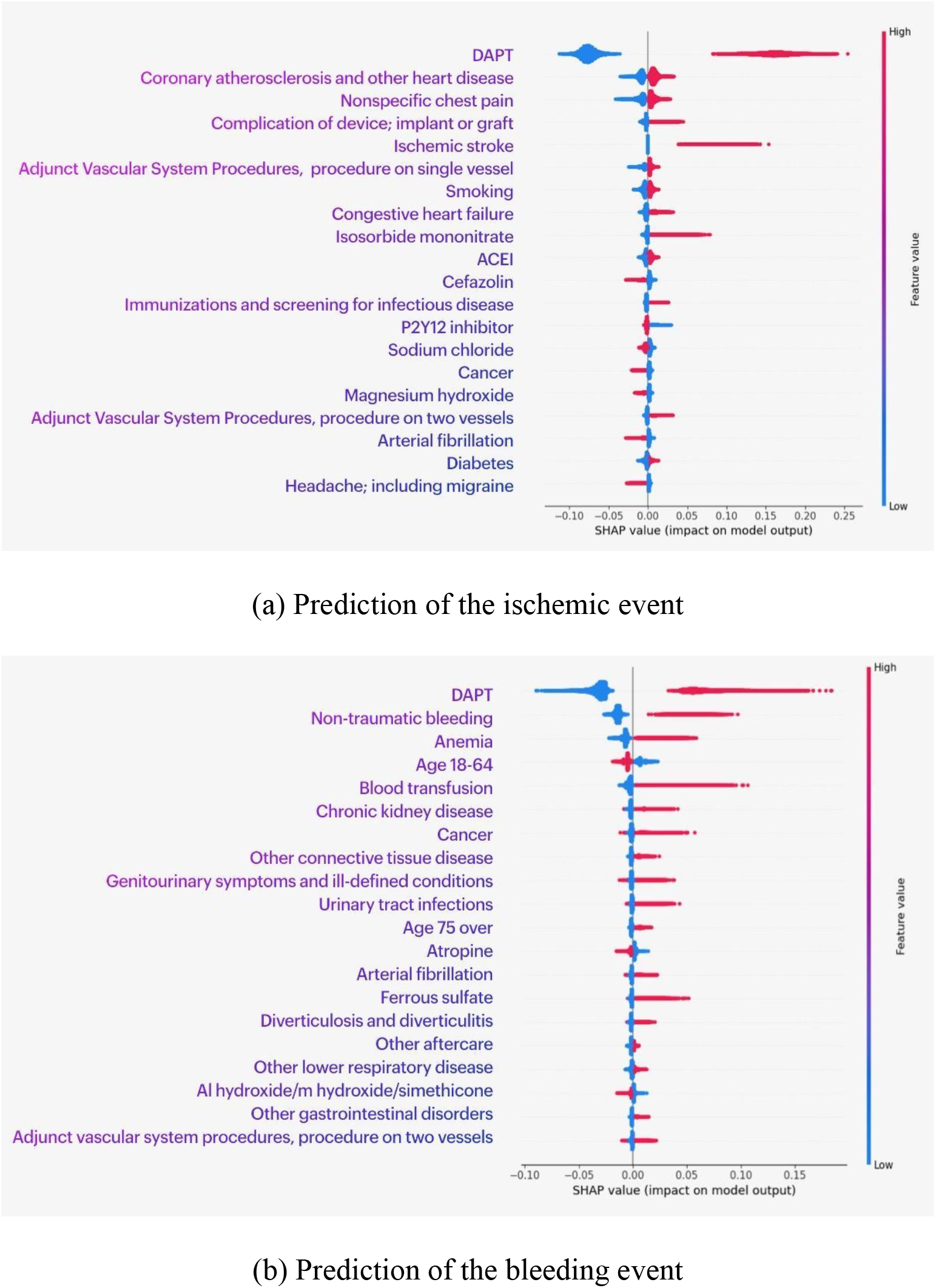
Top 20 features for ischemic and bleeding events prediction on the cohort level at 12m - 30m window

For the ischemic endpoint prediction, the highly contributing features include DAPT (DAPT, P2Y12 inhibitor), cardiovascular and cerebrovascular disease (coronary atherosclerosis and other heart diseases, ischemic stroke, nonspecific chest pain, congestive heart failure, atrial fibrillation), cardiovascular drugs (isosorbide mononitrate, ACEI), and possible risk factors (smoking, diabetes, and cancer) (Figure 4a).

For the bleeding endpoint prediction, the highly contributing features include DAPT, hematological disease (anemia, ferrous sulfate), prior bleeding (non-traumatic bleeding and blood transfusion), urinary system diseases (chronic kidney disease, urinary tract infections), and digestive disease (diverticulosis and diverticulitis, other gastrointestinal disorders) as well as possible risk factors (other connective tissue disease and cancer) (Figure 4b).

## Discussion

This study explored a novel dynamic prediction approach for ischemic and bleeding risk stratification after DES implantation leveraging cutting-edge AI models. To the best of our knowledge, this is the first effort that forecasts adverse endpoints after DES implantation with flexible and length-adjustable windows. The strengths of the study are summarized as follows: (1) We realized dynamic prediction in a variety of follow-up windows in comparison to the static data input and fixed prediction window of the existing scores (e.g., PRECISE-DAPT score, DAPT score). (2) Dynamic prediction algorithms can be integrated into computerized clinical monitoring systems, enabling clinicians to flexibly adjust treatment for patients. Counseling patients based on data available to cardiologists in different time windows could facilitate a better decision process regarding DAPT management. Healthcare under precision CV medicine will become a more integrated, dynamic system, in which patients are central stakeholders who contribute data and participate actively in shared decision making (42).

### Study Limitations

The results of our study should be considered in the context of several limitations: (1) The main limitation is the inherently incomplete nature of the EHR data (43). The incompleteness of medication-related data is evident in the Cerner data. For example, it is not uncommon that one patient had DAPT records at the index visit and another visit two years later, with missing information in between. It was thus difficult to judge whether the patient continued DAPT or not in the first year or the second year. To address this challenge, we stratified the patients into DAPT-continued or -discontinued groups based on a predefined time threshold. If the definition of DAPT continuation status was made on account of an inappropriate assumption, however, it could generate researcher-created bias. In addition, restricted by the structured EHR data, certain specific information was not available, including the metrics of the stent (diameter, length, and bifurcation), and cause of death (mainly cardiac death). This lack of certain information precluded further in-depth analysis. 2) The current study is dependent on standardized medical terminologies, including ICD-9, ICD-10, and CPT. We could not control the possibility of coding variability, which may lead to missed or incorrect reporting of certain conditions (44). For instance, there is no specific ICD code for stent thrombosis, which is an important component of the ischemic endpoint.

Our future efforts will address the limitations of missing or incomplete EHR data, especially the DAPT records therein. We will extend the databases to those that include more medication records, such as OneFlorida (45), Optum’s Clinformatics (46), and UT Physicians Clinical Data Warehouse (47). Further, we will integrate multi-modal data (such as unstructured clinical notes) as complementary information to enhance patient representation learning (48). To improve the model performance, we will integrate important features from existing scores to realize knowledge injection. Moreover, we will apply advanced EHR-tailored transformer-based approaches, such as Med-BERT (49). Overall, future studies should seek to overcome the existing limitations by using the datasets with higher quality and more advanced methodologies with superior efficacy.

## Conclusions

Optimum DAPT duration after DES implantation will likely benefit significantly from accurate prediction of the ischemic and bleeding risks. We developed a dynamic AI-empowered approach for forecasting adverse endpoints after PCI utilizing data available as treatment progresses. Our best performance model demonstrated robust discriminatory ability and meaningful risk stratification for both ischemic and bleeding endpoints. Dynamic clinical prediction models outperform static predictions made at a single time point. Our AI-DAPT score can potentially support clinical decision-making with respect to the optimum duration of antithrombotic treatment based on the patient’s personalized characteristics and increasingly available information, thus addressing the unmet need for precision patient-centered care.

## Perspectives

### Competency in patient care and procedural skills

The development of dynamic clinical prediction models represents an important methodological advance in the field of precision cardiovascular medicine because it improves the accuracy and potential clinical utility of such models. Dynamic prediction of ischemic and bleeding risks after DES implantation, given continued or discontinued DAPT, could guide clinicians to conduct more-precise risk evaluations and provide personalized antithrombotic therapy, which would greatly reduce patients’ life-threatening endpoints and mortality rates, ultimately improving their life quality.

### Translational outlook

Our results should be evaluated in further studies to better delineate the association between the DAPT duration and the possible risks of bleeding and ischemic endpoints.

## Supporting information

All Supplemental materials

## Data Availability

The data that support the findings of this study are available for licensing at Cerner Corporation

## Acknowledgments

We are grateful to the American Heart Association for providing the Precision Medicine Platform, which greatly supports our data processing and model development. We thank Dr. Zhen Zhou, M.D., for valuable discussions about code selection of standard terminologies for endpoint definitions. We thank Dr. Jianfu Li, Ph.D., for providing helpful technical support in the usage of GPU server and Visual Studio Code. We appreciate the great help of Sharon Lynn Bear, Ph.D. in language editing.

## Abbreviations

AI: artificial intelligence
CABG: coronary artery bypass graft
CKD: chronic kidney disease
CRUSADE: can rapid risk stratification of unstable angina patients suppress adverse outcomes with early implementation of the ACC/AHA guidelines
DAPT: dual antiplatelet therapy
PARIS: patterns of non-adherence to antiplatelet regimen in stented patients
PCI: percutaneous coronary intervention
PRECISE-DAPT: predicting bleeding complication in patients undergoing stent implantation and subsequent dual antiplatelet therapy

## References

1. Lawton JS, Tamis-Holland JE, Bangalore S, et al. 2021 ACC/AHA/SCAI guideline for coronary artery revascularization: A report of the American college of cardiology/American heart association joint committee on clinical practice guidelines. J. Am. Coll. Cardiol. 2021. Available at: http://dx.doi.org/10.1016/j.jacc.2021.09.006.

2. Bhatt DL. Percutaneous coronary intervention in 2018. JAMA 2018;319:2127–2128.

3. Buccheri D, Piraino D, Andolina G, Cortese B. Understanding and managing in-stent restenosis: a review of clinical data, from pathogenesis to treatment. J. Thorac. Dis. 2016;8:E1150–E1162.

4. Capodanno D, Alfonso F, Levine GN, Valgimigli M, Angiolillo DJ. ACC/AHA Versus ESC Guidelines on Dual Antiplatelet Therapy: JACC Guideline Comparison. J. Am. Coll. Cardiol. 2018;72:2915–2931.

5. Stefanini GG, Holmes DR Jr. Drug-eluting coronary-artery stents. N. Engl. J. Med. 2013;368:254–265.

6. Subherwal S, Bach RG, Chen AY, et al. Baseline risk of major bleeding in non-ST-segment-elevation myocardial infarction: the CRUSADE (Can Rapid risk stratification of Unstable angina patients Suppress ADverse outcomes with Early implementation of the ACC/AHA Guidelines) Bleeding Score. Circulation 2009;119:1873–1882.

7. Costa F, van Klaveren D, James S, et al. Derivation and validation of the predicting bleeding complications in patients undergoing stent implantation and subsequent dual antiplatelet therapy (PRECISE-DAPT) score: a pooled analysis of individual-patient datasets from clinical trials. Lancet 2017;389:1025–1034.

8. Baber U, Mehran R, Giustino G, et al. Coronary Thrombosis and Major Bleeding After PCI With Drug-Eluting Stents: Risk Scores From PARIS. J. Am. Coll. Cardiol. 2016;67:2224–2234.

9. Yeh RW, Secemsky EA, Kereiakes DJ, et al. Development and Validation of a Prediction Rule for Benefit and Harm of Dual Antiplatelet Therapy Beyond 1 Year After Percutaneous Coronary Intervention. JAMA 2016;315:1735–1749.

10. Jarrett D, Yoon J, van der Schaar M. Dynamic Prediction in Clinical Survival Analysis Using Temporal Convolutional Networks. IEEE J Biomed Health Inform 2020;24:424–436.

11. Xu Y, Liu X, Cao X, et al. Artificial intelligence: A powerful paradigm for scientific research. Innovation (N Y) 2021;2:100179.

12. Krittanawong C, Zhang H, Wang Z, Aydar M, Kitai T. Artificial Intelligence in Precision Cardiovascular Medicine. J. Am. Coll. Cardiol. 2017;69:2657–2664.

13. Al’Aref SJ, Singh G, Baskaran L, Metaxas D. Machine Learning in Cardiovascular Medicine. Academic Press; 2020.

14. Zack CJ, Senecal C, Kinar Y, et al. Leveraging Machine Learning Techniques to Forecast Patient Prognosis After Percutaneous Coronary Intervention. JACC Cardiovasc. Interv. 2019;12:1304–1311.

15. Mortazavi BJ, Bucholz EM, Desai NR, et al. Comparison of Machine Learning Methods With National Cardiovascular Data Registry Models for Prediction of Risk of Bleeding After Percutaneous Coronary Intervention. JAMA Network Open 2019;2:e196835. Available at: http://dx.doi.org/10.1001/jamanetworkopen.2019.6835.

16. Hsieh M-H, Lin S-Y, Lin C-L, et al. A fitting machine learning prediction model for short-term mortality following percutaneous catheterization intervention: a nationwide population-based study. Ann Transl Med 2019;7:732.

17. Rayfield C, Agasthi P, Mookadam F, et al. Machine Learning on High-Dimensional Data to Predict Bleeding Post Percutaneous Coronary Intervention. J. Invasive Cardiol. 2020;32:E122–E129.

18. Puget J-F. Feature engineering for deep learning. Inside Machine learning 2017. Available at: https://medium.com/inside-machine-learning/feature-engineering-for-deep-learning-2b1fc7605ace. Accessed July 26, 2021.

19. UTHealth. Cerner Health Facts. Available at: https://sbmi.uth.edu/sbmi-data-service/data-set/cerner/. Accessed December 16, 2019.

20. Mehran R, Rao SV, Bhatt DL, et al. Standardized bleeding definitions for cardiovascular clinical trials: a consensus report from the Bleeding Academic Research Consortium. Circulation 2011;123:2736–2747.

21. Sampedro-Gómez J, Dorado-Díaz PI, Vicente-Palacios V, et al. Machine Learning to Predict Stent Restenosis Based on Daily Demographic, Clinical, and Angiographic Characteristics. Can. J. Cardiol. 2020;36:1624–1632.

22. Elixhauser A. Concept: Elixhauser Comorbidity Index. Available at: http://mchp-appserv.cpe.umanitoba.ca/viewConcept.php?printer=Y&conceptID=1436. Accessed June 16, 2020.

23. AthenaHealth. Epocrates Web Drugs. Available at: https://online.epocrates.com/drugs. Accessed January 17, 2020.

24. Levine GN, Bates ER, Bittl JA, et al. 2016 ACC/AHA Guideline Focused Update on Duration of Dual Antiplatelet Therapy in Patients With Coronary Artery Disease: A Report of the American College of Cardiology/American Heart Association Task Force on Clinical Practice Guidelines: An Update of the 2011 ACCF/AHA/SCAI Guideline for Percutaneous Coronary Intervention, 2011 ACCF/AHA Guideline for Coronary Artery Bypass Graft Surgery, 2012 ACC/AHA/ACP/AATS/PCNA/SCAI/STS Guideline for the Diagnosis and Management of Patients With Stable Ischemic Heart Disease, 2013 ACCF/AHA Guideline for the Management of ST-Elevation Myocardial Infarction, 2014 AHA/ACC Guideline for the Management of Patients With Non-ST-Elevation Acute Coronary Syndromes, and 2014 ACC/AHA Guideline on Perioperative Cardiovascular Evaluation and Management of Patients Undergoing Noncardiac Surgery. Circulation 2016;134:e123–55.

25. Fan J, Ma X, Wu L, Zhang F, Yu X, Zeng W. Light Gradient Boosting Machine: An efficient soft computing model for estimating daily reference evapotranspiration with local and external meteorological data. Agric. Water Manage. 2019;225:105758.

26. Tolles J, Meurer WJ. Logistic Regression: Relating Patient Characteristics to Outcomes. JAMA 2016;316:533–534.

27. Svetnik V, Liaw A, Tong C, Culberson JC, Sheridan RP, Feuston BP. Random forest: a classification and regression tool for compound classification and QSAR modeling. J. Chem. Inf. Comput. Sci. 2003;43:1947–1958.

28. Cho K, van Merrienboer B, Gulcehre C, et al. Learning phrase representations using RNN encoder– decoder for statistical machine translation. In: Proceedings of the 2014 Conference on Empirical Methods in Natural Language Processing (EMNLP). Stroudsburg, PA, USA: Association for Computational Linguistics, 2014. Available at: http://dx.doi.org/10.3115/v1/d14-1179.

29. Choi E, Bahadori MT, Kulas JA, Schuetz A, Stewart WF, Sun J. RETAIN: An Interpretable Predictive Model for Healthcare using Reverse Time Attention Mechanism. arXiv [cs.LG] 2016. Available at: http://arxiv.org/abs/1608.05745.

30. Microsoft. LightGBM documentation. Available at: https://lightgbm.readthedocs.io/en/latest/index.html. Accessed February 17, 2021.

31. Rasmy L, Wu Y, Wang N, et al. A study of generalizability of recurrent neural network-based predictive models for heart failure onset risk using a large and heterogeneous EHR data set. J. Biomed. Inform. 2018;84:11–16.

32. Rasmy L, Xie Z, Zhi D, et al. pytorch_ehr: source codes based on PyTorch to analyze EHR. Available at: https://github.com/ZhiGroup/pytorch_ehr. Accessed December 17, 2020.

33. Rasmy L, Nigo M, Kannadath BS, et al. CovRNN—A recurrent neural network model for predicting outcomes of COVID-19 patients: model development and validation using EHR data. bioRxiv 2021:2021.09.27.21264121. Available at: https://www.medrxiv.org/content/10.1101/2021.09.27.21264121v1.full. Accessed December 17, 2021.

34. Akiba T, Sano S, Yanase T, Ohta T, Koyama M. Optuna: A Next-generation Hyperparameter Optimization Framework. In: Proceedings of the 25th ACM SIGKDD International Conference on Knowledge Discovery & Data Mining. KDD ‘19. New York, NY, USA: Association for Computing Machinery, 2019:2623–2631.

35. Kwok CS, Wong CW, Nagaraja V, Mamas MA. A systematic review of the studies that evaluate the performance of the DAPT score. Int. J. Clin. Pract. 2020;74:e13591.

36. Florencia M, Marcos V, Costabel JP, et al. PRECISE SCORE VALIDATION IN BUENOS AIRES 1 REGISTRY. Curr. Probl. Cardiol. 2022:101113.

37. Lundberg S. SHAP: A game theoretic approach to explain the output of any machine learning model. Github Available at: https://github.com/slundberg/shap. Accessed May 17, 2021.

38. Lundberg SM, Lee S-I. A unified approach to interpreting model predictions. In: Proceedings of the 31st international conference on neural information processing systems., 2017:4768–4777.

39. Casas P. How to interpret SHAP values in R (with code example!). Data Science Heroes Blog 2019. Available at: https://blog.datascienceheroes.com/how-to-interpret-shap-values-in-r/. Accessed February 5, 2022.

40. Rasmy L. Terminology_representation: Development of terminology representations for EHR prediction modeling. Github Available at: https://github.com/ZhiGroup/terminology_representation. Accessed January 8, 2022.

41. Agency for Healthcare Research and Quality. Clinical classifications software (CCS) for ICD-9-CM. Available at: https://www.hcup-us.ahrq.gov/toolssoftware/ccs/ccs.jsp. Accessed January 8, 2021.

42. Antman EM, Loscalzo J. Precision medicine in cardiology. Nat. Rev. Cardiol. 2016;13:591–602.

43. Hu Z, Melton GB, Arsoniadis EG, Wang Y, Kwaan MR, Simon GJ. Strategies for handling missing clinical data for automated surgical site infection detection from the electronic health record. J. Biomed. Inform. 2017;68:112–120.

44. Wijarnpreecha K, Li F, Xiang Y, et al. Nonselective beta-blockers are associated with a lower risk of hepatocellular carcinoma among cirrhotic patients in the United States. Aliment. Pharmacol. Ther. 2021;54:481–492.

45. University of Florida. OneFlorida Clinical Research Consortium. Available at: https://www.ctsi.ufl.edu/ctsa-consortium-projects/oneflorida/. Accessed March 19, 2021.

46. SBMI. Optum’s clinformatics® data mart. Available at: https://sbmi.uth.edu/sbmi-data-service/data-set/optum/. Accessed July 20, 2021.

47. UTHealth. UTPhysician clinical data warehouse. Available at: https://sbmi.uth.edu/sbmi-data-service/data-set/utphysician/. Accessed July 7, 2021.

48. Zhang D, Yin C, Zeng J, Yuan X, Zhang P. Combining structured and unstructured data for predictive models: a deep learning approach. BMC Med. Inform. Decis. Mak. 2020;20:280.

49. Rasmy L, Xiang Y, Xie Z, Tao C, Zhi D. Med-BERT: pretrained contextualized embeddings on large-scale structured electronic health records for disease prediction. NPJ Digit Med 2021;4:86.

