## Supplementary material for "AI-aided dynamic prediction of bleeding and ischemic risk after coronary stenting and subsequent DAPT": All Supplemental materials

Supplemental Appendix

### **1. Definitions of drug-eluting stent (DES) implantation**

**Supplementary Table 1.** Definitions of DES implantation

| **Name** | **Code type** | **Specific codes**^a^ |
| --- | --- | --- |
| Drug-eluting stent implantation | CPT4 | 92928, 92929, 92933, 92934 |
|  | HCPCS | C9600 - C9608 |
|  | ICD-9-PCS | 36.07 |
|  | ICD-10-PCS | 0270046, 0270056, 0270066, 0270076, 0270346, 0270356, 0270366, 0270376, 0270446, 0270456, 0270466, 0270476, 0271046, 0271056, 0271066, 0271076, 0271346, 0271356, 0271366, 0271376, 0271446, 0271456, 0271466, 0271476, 0272046, 0272056, 0272066, 0272076, 0272346, 0272356, 0272366, 0272376, 0272446, 0272456, 0272466, 0272476, 0273046, 0273056, 0273066, 0273076, 0273346, 0273356, 0273366, 0273376, 0273446, 0273456, 0273466, 0273476, 027004Z, 027005Z, 027006Z, 027007Z, 027034Z, 027035Z, 027036Z, 027037Z, 027044Z, 027045Z, 027046Z, 027047Z, 027104Z, 027105Z, 027106Z, 027107Z, 027134Z, 027135Z, 027136Z, 027137Z, 027144Z, 027145Z, 027146Z, 027147Z, 027204Z, 027205Z, 027206Z, 027207Z, 027234Z, 027235Z, 027236Z, 027237Z, 027244Z, 027245Z, 027246Z, 027247Z, 027304Z, 027305Z, 027306Z, 027307Z, 027334Z, 027335Z, 027336Z, 027337Z, 027344Z, 027345Z, 027346Z, 027347Z |

*Notes:*

Abbreviations: CPT4, Current Procedural Terminology, 4th Edition; HCPCS, Healthcare Common Procedure Coding System; ICD, International Classification of Diseases; PCS, Procedure Coding System

^a^If a medical code and all of its subclasses in the standard terminologies belong to the same clinical concept defined in our study, we keep the code at the highest level to reduce redundancy. This principle applies to all tables in this Supplementary Appendix.

### **2. Definitions of endpoints**

Ischemic endpoint was defined as the composite of acute ischemic heart disease, ischemic stroke, multiple drug-eluting stent implantation, and coronary artery bypass graft; and bleeding endpoint was defined as the composite of spontaneous bleeding and blood transfusion. See Supplementary Tables 2 and 3 for the specific ICD, CPT, and HCPCS codes.

**Supplementary Table 2.** Definitions of ischemic endpoints

| **Ischemic endpoints** | **Code type** | **Specific codes** |
| --- | --- | --- |
| Acute ischemic heart disease | ICD-9-CM | 410, 411, 427.5 |
|  | ICD-10-CM | I21, I22, I23, I24, I46 |
| Ischemic stroke | ICD-9-CM | 362.3, 433.x1, 434.x1, 435 |
|  | ICD-10-CM | G45, H34.1, I63, I67.81, I67.82 |
| Drug-eluting stent implantation^a^ | CPT4 | 92928, 92929, 92933, 92934 |
|  | HCPCS | C9600, C9601, C9602, C9603, C9604, C9605, C9606, C9607, C9608 |
|  | ICD-9-PCS | 36.07 |
|  | ICD-10-PCS | 0270046, 0270056, 0270066, 0270076, 0270346, 0270356, 0270366, 0270376, 0270446, 0270456, 0270466, 0270476, 0271046, 0271056, 0271066, 0271076, 0271346, 0271356, 0271366, 0271376, 0271446, 0271456, 0271466, 0271476, 0272046, 0272056, 0272066, 0272076, 0272346, 0272356, 0272366, 0272376, 0272446, 0272456, 0272466, 0272476, 0273046, 0273056, 0273066, 0273076, 0273346, 0273356, 0273366, 0273376, 0273446, 0273456, 0273466, 0273476, 027004Z, 027005Z, 027006Z, 027007Z, 027034Z, 027035Z, 027036Z, 027037Z, 027044Z, 027045Z, 027046Z, 027047Z, 027104Z, 027105Z, 027106Z, 027107Z, 027134Z, 027135Z, 027136Z, 027137Z, 027144Z, 027145Z, 027146Z, 027147Z, 027204Z, 027205Z, 027206Z, 027207Z, 027234Z, 027235Z, 027236Z, 027237Z, 027244Z, 027245Z, 027246Z, 027247Z, 027304Z, 027305Z, 027306Z, 027307Z, 027334Z, 027335Z, 027336Z, 027337Z, 027344Z, 027345Z, 027346Z, 027347Z |
| Coronary artery bypass graft | ICD-9-PCS | 36.1, 36.2, 36.3 |
|  | ICD-10-PCS | 0210083, 0210088, 0210089, 0210093, 0210098, 0210099, 0210344, 0210444, 0210483, 0210488, 0210489, 0210493, 0210498, 0210499, 0211083, 0211088, 0211089, 0211093, 0211098, 0211099, 0211344, 0211444, 0211483, 0211488, 0211489, 0211493, 0211498, 0211499, 0212083, 0212088, 0212089, 0212093, 0212098, 0212099, 0212344, 0212444, 0212483, 0212488, 0212489, 0212493, 0212498, 0212499, 0213083, 0213088, 0213089, 0213093, 0213098, 0213099, 0213344, 0213444, 0213483, 0213488, 0213489, 0213493, 0213498, 0213499, 021008C, 021008F, 021008W, 021009C, 021009F, 021009W, 02100A3, 02100A8, 02100A9, 02100AC, 02100AF, 02100AW, 02100J3, 02100J8, 02100J9, 02100JC, 02100JF, 02100JW, 02100K3, 02100K8, 02100K9, 02100KC, 02100KF, 02100KW, 02100Z3, 02100Z8, 02100Z9, 02100ZC, 02100ZF, 02103D4, 021048C, 021048F, 021048W, 021049C, 021049F, 021049W, 02104A3, 02104A8, 02104A9, 02104AC, 02104AF, 02104AW, 02104D4, 02104J3, 02104J8, 02104J9, 02104JC, 02104JF, 02104JW, 02104K3, 02104K8, 02104K9, 02104KC, 02104KF, 02104KW, 02104Z3, 02104Z8, 02104Z9, 02104ZC, 02104ZF, 021108C, 021108F, 021108W, 021109C, 021109F, 021109W, 02110A3, 02110A8, 02110A9, 02110AC, 02110AF, 02110AW, 02110J3, 02110J8, 02110J9, 02110JC, 02110JF, 02110JW, 02110K3, 02110K8, 02110K9, 02110KC, 02110KF, 02110KW, 02110Z3, 02110Z8, 02110Z9, 02110ZC, 02110ZF, 02113D4, 021148C, 021148F, 021148W, 021149C, 021149F, 021149W, 02114A3, 02114A8, 02114A9, 02114AC, 02114AF, 02114AW, 02114D4, 02114J3, 02114J8, 02114J9, 02114JC, 02114JF, 02114JW, 02114K3, 02114K8, 02114K9, 02114KC, 02114KF, 02114KW, 02114Z3, 02114Z8, 02114Z9, 02114ZC, 02114ZF, 021208C, 021208F, 021208W, 021209C, 021209F, 021209W, 02120A3, 02120A8, 02120A9, 02120AC, 02120AF, 02120AW, 02120J3, 02120J8, 02120J9, 02120JC, 02120JF, 02120JW, 02120K3, 02120K8, 02120K9, 02120KC, 02120KF, 02120KW, 02120Z3, 02120Z8, 02120Z9, 02120ZC, 02120ZF, 02123D4, 021248C, 021248F, 021248W, 021249C, 021249F, 021249W, 02124A3, 02124A8, 02124A9, 02124AC, 02124AF, 02124AW, 02124D4, 02124J3, 02124J8, 02124J9, 02124JC, 02124JF, 02124JW, 02124K3, 02124K8, 02124K9, 02124KC, 02124KF, 02124KW, 02124Z3, 02124Z8, 02124Z9, 02124ZC, 02124ZF, 021308C, 021308F, 021308W, 021309C, 021309F, 021309W, 02130A3, 02130A8, 02130A9, 02130AC, 02130AF, 02130AW, 02130J3, 02130J8, 02130J9, 02130JC, 02130JF, 02130JW, 02130K3, 02130K8, 02130K9, 02130KC, 02130KF, 02130KW, 02130Z3, 02130Z8, 02130Z9, 02130ZC, 02130ZF, 02133D4, 021348C, 021348F, 021348W, 021349C, 021349F, 021349W, 02134A3, 02134A8, 02134A9, 02134AC, 02134AF, 02134AW, 02134D4, 02134J3, 02134J8, 02134J9, 02134JC, 02134JF, 02134JW, 02134K3, 02134K8, 02134K9, 02134KC, 02134KF, 02134KW, 02134Z3, 02134Z8, 02134Z9, 02134ZC, 02134ZF |
|  | CPT4 | 33508, 33510, 33511, 33512, 33513, 33514, 33516, 33517, 33518, 33519, 33521, 33522, 33523, 33530, 33533, 33534, 33535, 33536, 33572 |

*Notes:*

^a^Multiple DES implantation after the first one (at the index visit) was regarded as the ischemic event.

**Supplementary Table 3.** Definitions of bleeding endpoints

| **Bleeding endpoints** | **Code type** | **Specific codes** |
| --- | --- | --- |
| Spontaneous bleeding | ICD-9-CM | 287.8, 287.9, 363.61, 363.62, 376.32, 377.42, 379.23, 423, 430, 431, 432, 456, 459, 530.21, 530.7, 530.82, 531, 532, 533, 534, 535.11, 535.21, 535.31, 535.41, 535.51, 535.61, 535.71, 537.83, 537.84, 562.02, 562.03, 562.12, 562.13, 569.3, 569.85, 578, 596.7, 599.7, 627.1, 719.1, 729.92, 784.8, 786.3 |
|  | ICD-10-CM | D62, D68.3, D69.8, D69.9, H05.23, H11.3, H21.0, H31.3, H35.6, H43.1, H47.02, H92.2, I23.0, I31.2, I60, I61, I62, I85.0, J94.2, K22.11, K22.6, K25.0, K25.2, K25.4, K25.6, K26.0, K26.2, K26.4, K26.6, K27.0, K27.2, K27.4, K27.6, K28.0, K28.2, K28.4, K28.6, K29.0, K31.811, K31.82, K55.21, K57.01, K57.11, K57.13, K57.21, K57.31, K57.33, K57.41, K57.51, K57.53, K57.81, K57.91, K57.93, K62.5, K66.1, K76.2, K92.0, K92.1, K92.2, M25.0, N02, N42.1, N92.0, N92.1, N92.3, N92.4, N93.0, N93.8, N93.9, N95.0, R04.0, R31, R58 |
| Blood transfusion | ICD-9-PCS | 99.0 |
|  | ICD-10-PCS | 30230H0, 30230H1, 30230K0, 30230K1, 30230L0, 30230L1, 30230M0, 30230M1, 30230N0, 30230N1, 30230P0, 30230P1, 30230R0, 30230R1, 30233H0, 30233H1, 30233K0, 30233K1, 30233L0, 30233L1, 30233M0, 30233M1, 30233N0, 30233N1, 30233P0, 30233P1, 30233R0, 30233R1, 30240H0, 30240H1, 30240K0, 30240K1, 30243H0, 30243H1 |
|  | CPT4 | 36430 |
|  | HCPCS | P9010, P9011, P9012, P9013, P9016, P9017, P9018, P9019, P9020, P9021, P9022, P9023, P9031, P9032, P9033, P9034, P9035, P9036, P9037, P9038, P9039, P9040, P9044, P9051, P9052, P9053, P9054, P9055, P9056, P9057, P9058, P9059, P9060, P9070, P9071, P9072, S2180, S3906, S9538 |

### **3. Definitions of comorbidities**

**Supplementary Table 4.** Definitions of comorbidities

| **Comorbidities and risk Factors** | **ICD-9-CM** | **ICD-10-PCS** |
| --- | --- | --- |
| Alcohol abuse | 265.2, 291, 303, 305.0, 357.5, 425.5, 535.3, 571.0, 571.1, 571.2, 571.3, 980, V11.3 | F10, G62.1, I42.6, K29.2, K70, T51, Z71.4 |
| Anemia | 280, 281, 282, 283, 284, 285 | D50, D51, D52, D53, D55, D56, D57, D58, D59, D60, D61, D62, D63, D64 |
| Atrial fibrillation | 427.31 | I48.1, I48.2, I48.91 |
| Cancer | 140, 141, 142, 143, 144, 145, 146, 147, 148, 149, 150, 151, 152, 153, 154, 155, 156, 157, 158, 159, 160, 161, 162, 163, 164, 165, 170, 171, 172, 173, 174, 175, 176, 179, 180, 181, 182, 183, 184, 185, 186, 187, 188, 189, 190, 191, 192, 193, 194, 195, 196, 197, 198, 199, 200, 201, 202, 203, 204, 205, 206, 207, 208, 209, 230, 231, 232, 233, 234, 511.81 | C00, C01, C02, C03, C04, C05, C06, C07, C08, C09, C10, C11, C12, C13, C14, C15, C16, C17, C18, C19, C20, C21, C22, C23, C24, C25, C26, C30, C31, C32, C33, C34, C37, C38, C39, C40, C41, C43, C44, C45, C46, C47, C48, C49, C50, C51, C52, C53, C54, C55, C56, C57, C58, C60, C61, C62, C63, C64, C65, C66, C67, C68, C69, C70, C71, C72, C73, C74, C75, C76, C77, C78, C79, C7A.0, C80, C81, C82, C83, C84, C85, C86, C88, C90, C91, C92, C93, C94, C95, C96, D00, D01, D02, D03, D04, D05, D06, D07, D09, J91.0, O9A, R97.21, V10, V71.1, Z08, Z40.0, Z85 |
| Chronic kidney disease | 285.21, 403, 404, 585 | D63.1, N18, I12, I13 |
| Congestive heart failure | 398.91, 402.01, 402.11, 402.91, 404.01, 404.03, 404.11, 404.13, 404.91, 404.93, 425.4, 425.5, 425.7, 425.8, 425.9, 428 | I09.9, I11.0, I13.0, I13.2, I25.5, I42.0, I42.5, I42.6, I42.7, I42.8, I42.9, I43, I50, P29.0 |
| Diabetes mellitus | 249, 250, 648 | E08, E09, E10, E11, E13, O24 |
| Dyslipidemia | 272 | E78 |
| Hypertension | 401, 402, 403, 404, 405 | I10, I11, I12, I13, I15, I16 |
| Liver disease | 070, 121.3, 155.0, 155.2, 570, 571, 572, 573, 456.0, 456.1, 456.2, V42.7 | B15, B16, B17, B18, B19, B66.3, C22.0, C22.2, C22.3, C22.4, C22.7, C22.8, C22.9, K70, K71, K72, K73, K74, K75, K76, K77, Z94.4, I85, I86.4 |
| Peripheral vascular disease | 093.0, 415, 416.2, 437.3, 433, 440, 441, 443, 444, 445, 447, 557.1, 557.9, V43.4 | I26, I27.82, I63.0, I63.1, I70, I71, I73, I74, I75, I77.1, I77.3, I82.0, K55.1, K55.8, K55.9, I79.0, Z95.8, Z95.9 |
| Smoking-related disease | 305.1, 649, 989.84 | F17.200, F17.201, F17.210, F17.211, F17.220, F17.221, F17.290, F17.291, V15.82, Z87.891, Z72.0, Z71.6, Z87.891, T65.2 |
| Stroke^1,2^ | 362.3, 430, 431, 432, 433.x1, 434.x1, 435, 436, 437, 438 | G45, G46, H34.1, I60, I61, I63, I67.81, I67.82, I67.89 |

### **4. Definitions of DAPT and comedications**

DAPT definition: aspirin + a P2Y12 inhibitor (clopidogrel, prasugrel, ticagrelor)

For comedication definitions, see Supplementary Table 5 below.

**Supplementary Table 5.** Definitions of comedications

| **Therapeutic or Pharmacological Class** | **Generic name** |
| --- | --- |
| Anticoagulant | warfarin, apixaban, betrixaban, dabigatran, edoxaban, rivaroxaban |
| ACEI | amlodipine-benazepril, benazepril, captopril, enalapril, enalaprilat, fosinopril, lisinopril, moexipril, perindopril erbumine, quinapril, ramipril, trandolapril, trandolapril-verapamil |
| ARB | amlodipine-olmesartan, amlodipine-telmisartan, amlodipine-valsartan, azilsartan, candesartan, candesartan cilexetil, eprosartan, irbesartan, losartan, nebivolol-valsartan, olmesartan, sacubitril-valsartan, telmisartan, valsartan |
| Beta-blocker | acebutolol, alprenolol, atenolol, betaxolol, bisoprolol, carteolol, carvedilol, esmolol, labetalol, levobunolol, metoprolol, nadolol, nebivolol, nebivolol-valsartan, oxprenolol, penbutolol, pindolol, propranolol, sotalol, timolol |
| Calcium antagonist | aliskiren-amlodipine, aliskiren/amlodipine/hydrochlorothiazide, amlodipine, amlodipine-atorvastatin, amlodipine-benazepril, amlodipine-celecoxib, amlodipine-olmesartan, amlodipine-perindopril, amlodipine-telmisartan, amlodipine-valsartan, amlodipine/hydrochlorothiazide/olmesartan, clevidipine, diltiazem, felodipine, isradipine, levamlodipine, nicardipine, nifedipine, nimodipine, nisoldipine, trandolapril-verapamil, verapamil |
| NSAID | amlodipine-celecoxib, celecoxib, diclofenac, diclofenac topical, diclofenac-misoprostol, esomeprazole-naproxen, etodolac, famotidine-ibuprofen, fenoprofen, flurbiprofen, ibuprofen, indomethacin, ketoprofen, ketorolac, meclofenamate, mefenamic acid, meloxicam, nabumetone, naproxen, naproxen-sumatriptan, oxaprozin, piroxicam, sulindac, tolmetin |
| PPI | amoxicillin/clarithromycin/lansoprazole, amoxicillin/clarithromycin/omeprazole, dexlansoprazole, esomeprazole, lansoprazole, omeprazole, omeprazole-sodium bicarbonate, pantoprazole, rabeprazole |
| Statins | amlodipine-atorvastatin, atorvastatin, atorvastatin-ezetimibe, cerivastatin, ezetimibe-simvastatin, fluvastatin, lovastatin, pitavastatin, pravastatin, rosuvastatin, simvastatin, simvastatin-sitagliptin |

*Notes:*

Abbreviations: ACEI, angiotensin-converting enzyme inhibitor; ARB, angiotensin receptor blocker; NSAID, non-steroidal anti-inflammatory drug; PPI, proton pump inhibitor

### **5. Cohort number, DAPT treatment, and event case/control distribution**

**Supplementary Table 6.** Cohort distribution

| **Prediction window** | **Total cohort** | **With DAPT (%)** | **Ischemic event** | | **Bleeding event** | |
| --- | --- | --- | --- | --- | --- | --- |
|  |  |  | **Case** | **Control** | **Case** | **Control** |
| 12–15 m | 81,594 | 26,832 (32.9%) | 3,139 | 78,455 | 1,934 | 79,660 |
| 15–18 m | 76,658 | 25,610 (33.4%) | 2,564 | 74,094 | 1,702 | 74,956 |
| 18–21 m | 72,256 | 24,162 (33.4%)) | 2,284 | 69,972 | 1,536 | 70,720 |
| 21–24 m | 68,086 | 22,624 (33.2%) | 2,228 | 65,858 | 1,476 | 66,610 |
| 12–18 m | 81,594 | 26,832 (32.9%) | 5,293 | 76,301 | 3,362 | 78,232 |
| 18–24 m | 72,256 | 24,162 (33.4%) | 4,217 | 68,039 | 2,816 | 69,440 |
| 12–24 m | 81,594 | 26,832 (32.9%) | 8,678 | 72,916 | 5,699 | 75,895 |
| 12–30 m | 81,594 | 26,832 (32.9%) | 11,039 | 70,555 | 7,402 | 74,192 |

### **6. Conversion of Cox C-statistic to AUC**

Pseudocodes:

\# feed data to Cox and get the estimator for both c statistic and AUC

Y_cStatistic_test = data[['event_label', 'time_to_event']]

estimator = CoxPHSurvivalAnalysis(alpha=0.0001).fit(X_train, Y_c_statistic_test)

\# C-statistic calculation

c_statistic_score = estimator.score(X_test, Y_cStatistic_test)

\# AUC calculation

Y_auc_test = data[['event_label']]

risk_score = estimator.predict(X_test)

y_score = np.array(risk_score)

AUC = roc_auc_score(Y_auc_test, y_score)

### **7. Hyperparameter search space of AI models**

**Supplementary Table 7.** Hyperparameter search space of AI models

| **AI model** | **Hyperparameter search space** |
| --- | --- |
| LGBM | lambda_l1: 1e-8, 10.0; lambda_l2: 1e-8, 10.0; num_leaves: 2, 256; feature_fraction: 0.4, 1.0; bagging_fraction: 0.4, 1.0; bagging_freq: 1, 7; min_child_samples: 5, 100 |
| LR | logreg_c: e^-10^, e^10^ |
| RF | n_estimators: 2, 100; max_depth: 2, 30 |
| BiGRU | lr: 10^-5^, 10^-1^; l2: 10^-5^, 10^-2^, eps: 10^-5^, 10^-3^, embed_dim: 2^6^, 2^8^, hidden_size: 2^6^, 2^8^, optimizer: Adam, Adagrad, Adamax, Adadelta |
| RETAIN | lr: 10^-5^, 10^-1^; l2: 10^-5^, 10^-2^, eps: 10^-5^, 10^-3^, embed_dim: 2^6^, 2^8^, hidden_size: 2^6^, 2^8^, optimizer: Adam, Adagrad, Adamax, Adadelta |

Abbreviations: LR, logistic regression; RF, random forest; LGBM, light gradient boosting machine; BiGRU, bidirectional gated recurrent unit; RETAIN, REverse Time AttentIoN model

**References**

1.	Liu S, Chan W-S, Ray JG, Kramer MS, Joseph KS, for the Canadian Perinatal Surveillance System (Public Health Agency of Canada). Stroke and cerebrovascular disease in pregnancy. *Stroke*. 2019;50(1):13-20.

2.	Kokotailo RA, Hill MD. Coding of stroke and stroke risk factors using international classification of diseases, revisions 9 and 10. *Stroke*. 2005;36(8):1776-1781.
